# Community level variability in Bronx COVID-19 hospitalizations associated with differing viral variant adaptive strategies during the second year of the pandemic

**DOI:** 10.1101/2024.03.05.24303791

**Authors:** Ryan Forster, Anthony Griffen, Johanna Daily, Libusha Kelly

**Author notes:** Corresponding authors (L.K.); (J.P.D.).

## Abstract

The Bronx, New York, exhibited unique peaks in the number of COVID-19 cases and hospitalizations compared to national trends. To determine which features of the SARS-CoV-2 virus might underpin this local disease epidemiology, we conducted a comprehensive analysis of the genomic epidemiology of the four dominant strains of SARS-CoV-2 (Alpha, Iota, Delta and Omicron) responsible for COVID-19 cases in the Bronx between March 2020 and January 2023. Genomic analysis revealed similar viral fitness for Alpha and Iota variants in the Bronx compared to nationwide data. However, Delta and Omicron variants had increased fitness within the borough. While the transmission dynamics of most variants in the Bronx corresponded with mutational fitness-based predictions of transmissibility, the Delta variant presented as an exception. Epidemiological modeling confirms Delta’s advantages of higher transmissibility, and suggested pre-existing immunity within the community counteracted Delta virulence, contributing to unexpectedly low Bronx hospitalizations compared to preceding strains. There were few novel T-cell epitope mutations in Delta compared to Iota which suggests Delta had fewer immune escape mechanisms to subvert pre-existing immunity within the Bronx. The combination of epidemiological models and quantifying amino acid changes in T-cell and antibody epitopes also revealed an evolutionary trade-off between Alpha’s higher transmissibility and Iota’s immune evasion, potentially explaining why the Bronx Iota variant remained dominant despite the introduction of the nationwide dominant Alpha variant. Together, our study demonstrates that localized analyses and integration of orthogonal community-level datasets can provide key insights into the mechanisms of transmission and immunity patterns associated with regional COVID-19 incidence and disease severity that may be missed when analyzing broader datasets.

**One sentence summary:** A reduction in COVID-19 cases and hospitalizations during the Delta variant’s dominance are preceded with the prevalence of the immune-evading Iota variant over the globally dominant, more transmissible Alpha variant amongst the Bronx, indicating community-level variability associated with differing viral variant adaptive strategies.

## Background

Bronx, New York, a borough of New York City, was an early epicenter of SARS-CoV-2 transmission during the early period of the pandemic (CDC 2022). The cumulative rate of confirmed cases of COVID-19 in the Bronx as of September 11th, 2022 was 11,495 cases/100,000 individuals. The Bronx has 1,472,654 residents, is a large, densely populated population, approximately 34,920 people / sq. mile, and has an ethnically and racially diverse population. It is the third largest county by population density and the 13th in overall population within the U.S. Of the New York City boroughs, the Bronx had the highest hospitalization and death rate due to COVID-19: 1,752 hospitalizations/100,000 individuals and 470 deaths/100,000 individuals over the three years of the pandemic. Several factors likely contributed to these high rates. Residents of the Bronx experience high rates of asthma and other chronic respiratory ailments, which are known risk factors for severe COVID-19 (CDC 2020 & United States Census Bureau 2021). Furthermore, certain demographic groups are severely impacted by COVID-19, including Black/African American, Native American, Asian, or Hispanic/Latino individuals, as well as those of low socio-economic status. More than 50% of the Bronx population are Black/African American or Hispanic/Latino descent (CDC 2020) and 24% of residents are below the poverty line. Taken together, the Bronx is home to a uniquely vulnerable population to SARS-CoV-2 infection and severe COVID-19.

To understand the local determinants of SARS-CoV2 infection, we previously investigated the viral diversity of SARS-CoV-2 in the Bronx during the first year of the pandemic and found that SARS-CoV-2 viral diversity in the Bronx matched global trends, except for the emergence of a Bronx-originating lineage (Fels et al. 2022). Chronic infection of immunocompromised individuals by SARS-CoV-2 has been observed in the Bronx previously (Fels et al. 2022) and has been linked to an accelerated mutation rate and is proposed to be a mechanism responsible for the emergence of variants of concern (Chaguza et al. 2023 & Kemp et al. 2021), making local surveillance efforts in vulnerable patient populations such as the Bronx critical. Genomic surveillance during the second year of the pandemic conducted in New York State (NYS) identified a unique viral lineage: B.1.526, the New York-originating Iota variant of interest (VOI). Iota was the dominating lineage in NYS in contrast to the globally dominating lineage at the time: B.1.1.7, the Alpha variant of concern (VOC). The lineages B.1.1.7 and B.1.526 were not preferentially represented in vaccine breakthrough patients, compared to unvaccinated patients infected during the same sampling period, implying that the SARS-Cov-2 vaccine-induced immunity against each lineage was similar (Duerr et al. 2021). Despite this observation, estimates of transmissibility suggested Alpha was more transmissible than Iota, however these studies were either limited to the genomes selected or investigated over larger geographic regions (Dellicour et al. 2023, Petrone Me et al. 2022 & Annavajhala et al. 2021). As the pandemic has progressed, new VOCs have emerged, and a greater understanding of how immune escape mutations and single nucleotide positions facilitate transmission has developed (Wright et al. 2022 & Obermeyer et al. 2022).

We observed that the Bronx had unique peaks in COVID-19 cases, hospitalizations, and dynamics of VOI and VOC compared to national data. To determine which features of the SARS-CoV-2 virus and population health data might explain these differences, we characterized the longitudinal genomic epidemiology of SARS-CoV-2 in the Bronx from March 2020 until January 2023. We quantified viral dynamics, mutational fitness, antibody escape, T-cell epitope changes, and transmissibility and analyzed how these are associated with large-scale population health data. Our studies provide a comprehensive assessment of how different viral variants interacted with the Bronx population over time. This multifaceted approach reveals intricate interactions between SARS-CoV-2 variants and the immune landscape in the Bronx, highlighting the critical role of local surveillance for tailoring regional public health strategies and interventions against constantly evolving pandemic dynamics.

## Results

### Bronx deviates from U.S.A aggregate data during the second year of the SARS-CoV-2 pandemic in COVID-19 case and hospitalization rates

Trends in case rates and hospitalizations resulting from SARS-CoV2 infection significantly differed in the Bronx when compared with US aggregate data (**Fig. 1A & 1B**). Bronx case rates, measured per 100,000 individuals defined by positive SARS-CoV2 PCR tests, show five peaks over the three years of the pandemic. Wastewater PCR testing for the SARS-CoV-2 N gene complements patient COVID-19 rates and reveals a community viral load mirroring major trends in case rates. Four out of five peaks in Bronx’s COVID-19 cases surpass the Bronx’s average case rate for the entire pandemic, except for the summer of 2021, where the peak falls below the 99% confidence interval. However, the community viral load measured via wastewater remains above the 99% confidence interval for the same time period (**Fig. 1A**). Correspondingly, Bronx hospitalizations due to COVID-19 align with the observed trends in case rates.

**Figure 1:**
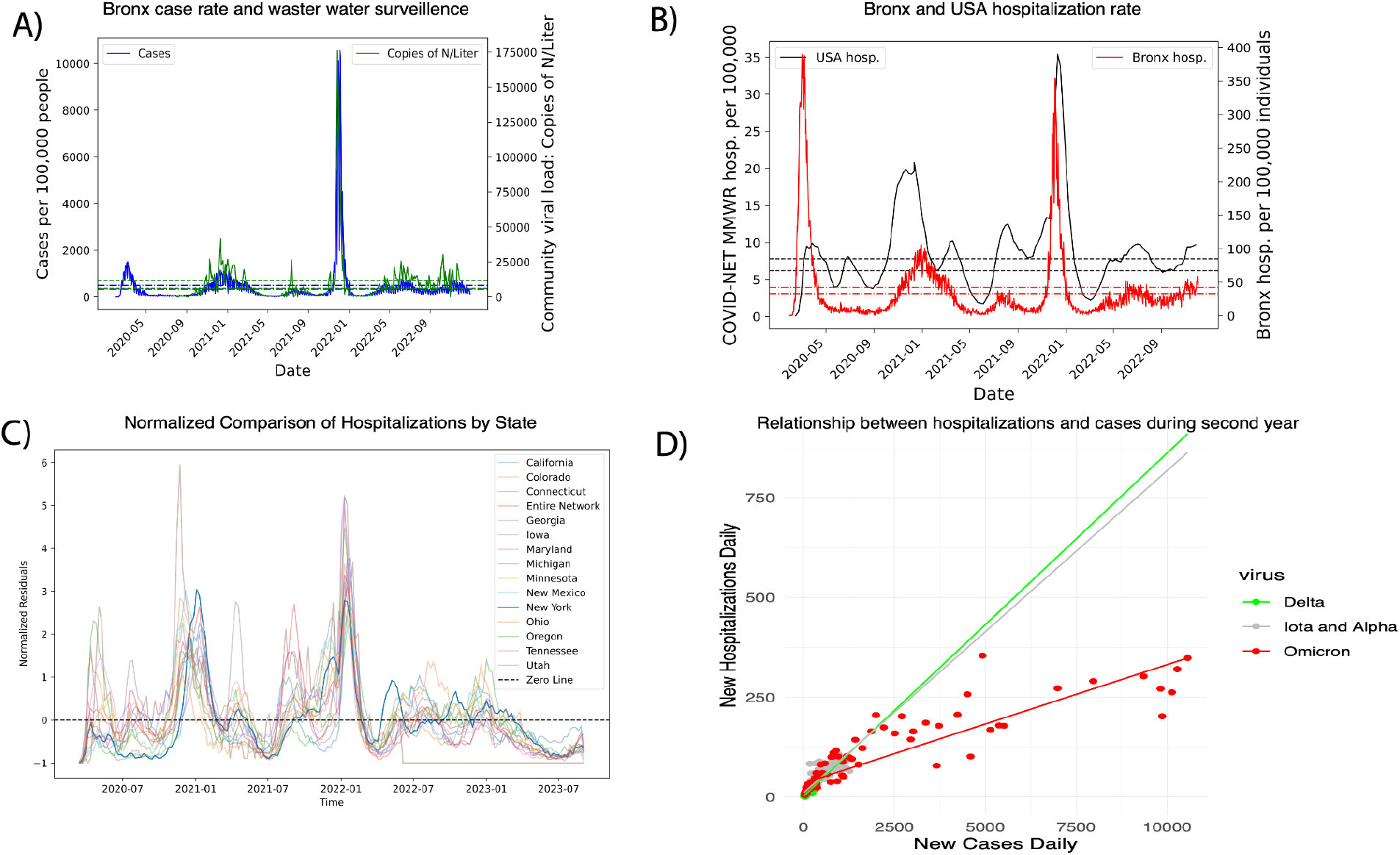
Bronx COVID-19 hospitalizations on average are below the U.S.A aggregate during year two of the pandemic, and the hospitalizations per case during the period of Delta VOC dominance are equivalent to the hospitalizations per case before it arrived in the Bronx. **A)** A plot of SARS-CoV-2 PCR diagnosed cases per 100,000 individuals in the Bronx compared with wastewater quantification of SARS-CoV-2 community load via qPCR of the nucleocapsid protein. **B)** A plot of Bronx and U.S.A hospitalizations due to COVID-19 per 100,000 individuals over time. The dotted lines indicate the 99% confidence interval values for a time series average any part of the time series above it is higher than average. **C)** Normalized COVID-19 hospitalizations by state were plotted and directly compared, variability above and below the zero line indicates different behavior during a given period of time. **D)** A robust linear model of the relationship between cases and hospitalizations during the second year and accounted for the statistical interaction between dominating VOC and cases with no significant difference in new hospitalizations per new case during Delta VOC dominance, p-value = 0.65.

Notably, Bronx hospitalizations are below average for the summer of 2021, in contrast to the U.S.A aggregate, which remains above average during the same period (**Fig. 1B**). The Bronx peaks in winter cases had statistically significant autocorrelation in hospitalizations and cases, with the strongest correlation between the original introduction of SARS-CoV-2 and the winter of 2021 (**Supp. Fig. 1**). To understand variability nationwide during the three years of the pnademic, we furhter analyzed state-level data across the U.S.. Splitting by state and normalizing by hospitalizations, variations above and below the state averages are evident during the summer of 2021, with consistent trends observed during the winter peaks and inconsistency during the summer months (**Fig. 1C**). The summer of 2021 is associated with the arrival of the Delta VOC, which is considered a more virulent strain (Carabelli et al. 2023 & Dellicour et al. 2023), yet there were lower cases and hospitalizations within the Bronx (**Fig. 1A & 1B**). To explore the relationship between cases and hospitalizations during the second year we employed a robust linear model, incorporating cases, hospitalizations, and an interaction term for the dominating VOC. The analysis revealed no statistical difference in the slope of the regressions between the periods dominated by (Rambaut et al. 2020) Delta and Iota during the second year (**Fig. 1D**).

### Genomic analysis demonstrates that Bronx Alpha and Iota VOC had equivalent fitness and Delta had increased mutational fitness

To further investigate the trends in VOC and VOI in the Bronx over the entirety of the pandemic, the Nextstrain pipeline (Hadfield et al.) was implemented to examine sequence variations.

SARS-CoV-2 isolates in the Bronx had an observed rate of 25 nucleotide substitutions per year, slightly below the worldwide average of 28 nucleotide substitutions per year (source: “Auspice” 2023) from March 2020 to October 2023. Of 12 VOCs identified in the Bronx, only four are well-represented in the sampling process. These are, in chronological order of appearance in the Bronx: Iota, Alpha, Delta, and Omicron, each with a higher number of mutations compared to the preceding VOC (**Fig. 2A**). To assess the fitness of each virus based on their mutations, the Nextstrain pipeline employs the PyR_0_ pre-trained machine learning predictor. This hierarchical Bayesian multinomial logistic regression model, quantifies relative fitness (exponential growth rate) of each virus. Despite the higher mutation count in Bronx Alpha sequences compared to Iota, the mutational fitness of Alpha and Iota was found to be identical.

**Figure 2:**
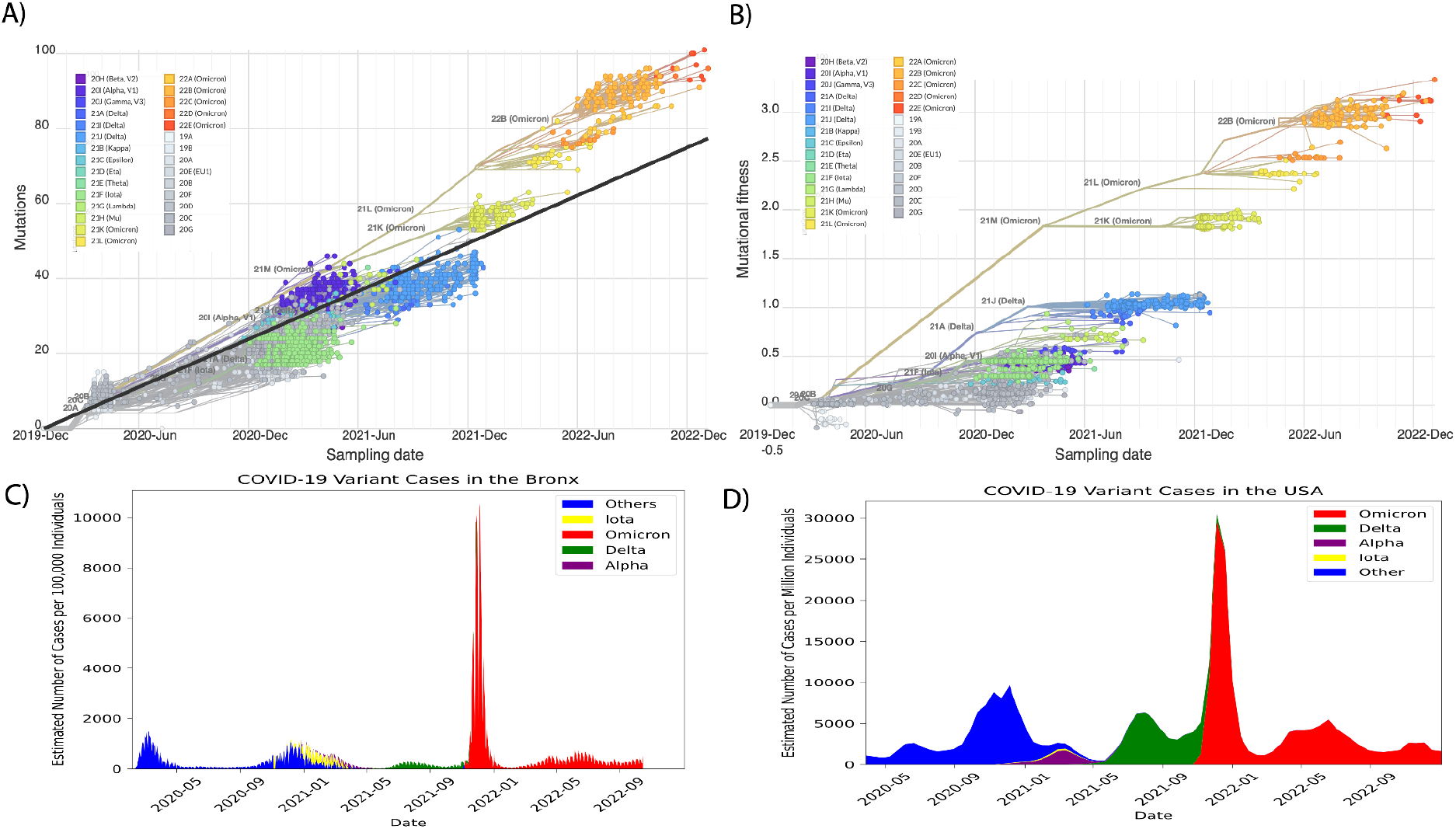
Alpha and Iota have equivalent mutational fitness and despite having a higher fitness, the estimated proportion of Delta cases are less than Iota cases in the Bronx. **A)** Mutation rate regression of 2,792 viruses sequenced in the Bronx, points are connected by phylogenetic order. On the x-axis is sampling date and on the y-axis is the number of nucleotide mutations. The mutation rate is 25 nucleotide substitutions a year relative to the Wuhan reference. **B)** A similar regression plot connected by phylogeny, using mutational fitness in place of the number of mutations on y-axis. Mutational fitness, or the relative combination of mutations that contribute to continued transmission is inferred from a hierarchical Bayesian multinomial logistic regression model: PyRo as part of the NextStrain pipeline. In plots A and B, coloring depicts the Nextrain clade each virus belongs to. Estimated proportion of cases by major VOC in **C)** the Bronx and **D)** the U.S.A colored by major VOC/VOI.

Conversely, Delta and Omicron exhibited an expected increase in fitness. The preceding first-year lineages had the lowest mutational fitness (**Fig. 2B**). Before the arrival of Iota in the Bronx, there was a higher number of Pangolin lineages (Rambaut et al. 2020) compared to any other point in the pandemic, but a lower number of overall mutations and non-synonymous mutations per virus (**Supp. Fig. 2 & 3, Fig. 2A**). The proportional distribution of cases, as estimated from limited sampling, indicates that Iota dominated and continued to do so even after the introduction of Alpha in the Bronx, in contrast with the broader U.S.A trend where Alpha was the predominant VOC. The decline in both Alpha and Iota cases, coupled with an overall reduction in lineage diversity, preceded the arrival of Delta, coinciding with a peak in cases during the summer of 2021 (**Fig. 2C & 2D**). Notably, this peak correlated with a general decrease in cases and hospitalizations (**Fig. 1A & 1B**).

### Estimation of transmission and recovery parameters of Bronx Iota, Alpha and Delta

We then conducted modeling of Bronx variant case data in year two of the pandemic, specifically focusing on the Iota, Alpha, and Delta variants, which exhibited distinct differences compared to the overall U.S.A aggregate. To mitigate the impact of noise in the case data, a monthly trend was established by smoothing the estimated case data specifically for the second year of the pandemic, employing a kernel density function to estimate cases. The smoothed Delta cases suggest that it arrived in the Bronx as early as May 2021, a month before it was detected (**Fig. 3A**). The doubling time for the smoothed cases to reach their peak cases was 14 days for Delta, 35 days for Alpha, and 40 days for Iota under an exponential growth model (R^2^ = 0.87 - 0.98, p-values < 2.2^-16^). For a comparative analysis of transmission dynamics using the SIR model, the parameters λ (representing the rate of new cases per day) and R_0_ (the number of secondary infections per primary infection) were derived from fitting to smoothed case data. The parameter λ is defined as the rate of new cases per day and is directly proportional to the slope of the daily cases regressed against cumulative cases. The linear regression analysis of daily cases against cumulative cases revealed a steeper slope for Delta cases (R^2^ = 0.9673, p-value < 2.2^-16^), while Iota (R^2^ = 0.981, p-value < 2.2^-16^) and Alpha (R^2^ = 0.9244, p-value < 2.2^-16^) exhibited similar slopes and better fit to data than the logistic growth model (**Fig. 3B & Table 1**). Regarding R_0_, which is under the SIR model, representing the ratio of the probability of infection (**β**) to the probability of recovery (**γ**), a dynamic and stochastic SIR model was employed to estimate β, γ, and R_0_. The R_0_ was found to be equivalent between Alpha and Iota, but higher in the case of Delta. Alpha demonstrated an equivalent β compared to Delta, although Alpha had the highest γ. Conversely, Delta and Iota exhibited lower γ than Alpha. The results were consistent between the dynamic model and the stochastic model and all models’ results were statistically significant with p-values < 2.2^-16^ . The dynamic model had a better fit to smoothed cases for Iota (R^2^ = 0.9966), compared with Alpha (R^2^ = 0.861) and Delta (R^2^ = 0.8471), however the stochastic model of Delta (R^2^ = 0.9488) had a better fit than Iota (R^2^ = 0.866) and Alpha (R^2^ = 0.8451) (**Fig. 3C, 3D & Table 1**). Overall the SIR models agree that there was a greater probability of recovery from infection after the introduction of Iota.

**Table 1:** Parameter values for transmission dynamics λ, cβ, γ, and R_0_ are listed for each model and virus, indicating higher transmissibility of Delta VOC during the second year.

|  | Iota | Alpha | Delta |
| --- | --- | --- | --- |
| **Force of infection regression** | $\lambda = 0.0164587$ | $\lambda = 0.0171035$ | $\lambda = 0.0388012$ |
| **Dynamic SIR** | $c\beta = 0.5161696$ | $c\beta = 1.280685$ | $c\beta = 1.278657$ |
| | $\gamma = 0.4553649$ | $\gamma = 1.118557$ | $\gamma = 0.8793897$ |
| | $R_0 = 1.13353$ | $R_0 = 1.118557$ | $R_0 = 1.454028$ |
| **Stochastic SIR** | $c\beta = 0.7594699$ | $c\beta = 1.060927$ | $c\beta = 1.283294$ |
| | $\gamma = 0.6666529$ | $\gamma = 0.9501149$ | $\gamma = 0.8768929$ |
| | $R_0 = 1.139228$ | $R_0 = 1.121587$ | $R_0 = 1.463456$ |

**Figure 3:**
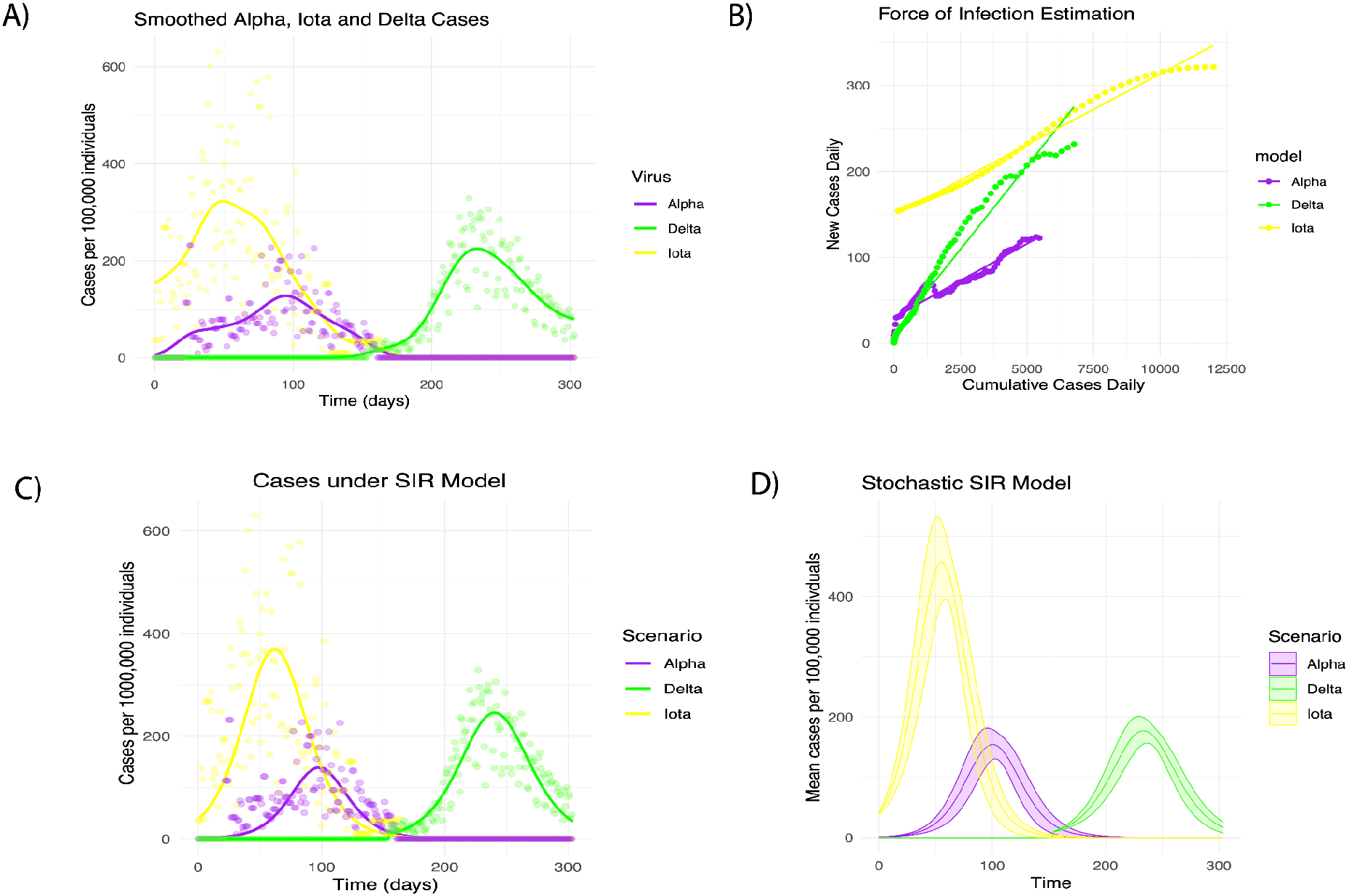
Fitting estimated case data of major VOC during the second year to estimate the force of infection (λ), transmission rate (β), and recovery rate (γ). **A)** A plot of the kernel density smoothed cases of Iota, Alpha and Delta variants. **B)** A plot of the cumulative cases regressed with the daily cases over time. **C)** A plot visualizing the optimized dynamic SIR models case trajectory for the three variants. **D)** The same plot in C visualizing the mean trajectory and standard deviation from the optimized stochastic differential SIR model in place of the dynamic model.

### Bronx Delta VOC has equivalent T-cell epitope variability to Iota, while Bronx Alpha epitope mutations were more similar to ancestral clades

To understand viral adaptation to the immune system, we incorporated population-level data on the number of mutations in immune epitopes, and viral mutations associated with monoclonal antibody resistance and antiviral treatments sourced from the COGS-UK database. Our findings (**Fig. 4A**), revealed most immune epitope changes observed in the database were also observed in the Bronx, with the majority situated in the spike protein. Omicron exhibited the widest range of T-cell epitope mutations, while Alpha had the narrowest range of epitope variation, with Iota surpassing Delta in this aspect. Regarding mutations affecting antibodies, including epitope changes or alterations that impact neutralization activity, Omicron had the most substantial number of mutations that would impact putative protective antibodies, outnumbering all other variants. Delta, while trailing behind Omicron, exhibited a slightly higher population count of antibody-impacting mutations compared to Iota, with Alpha recording the lowest count per virus and across the population (**Fig. 4A & Supp. Fig. 4**). Over the timeframe spanning March 2020 to January 2023, a consistent upward trajectory was observed in average-monthly amino acid changes across all viral genes, reaching a peak of 50 changes. T-cell epitope mutations and those conferring resistance to monoclonal antibodies followed a similar trend but with lower average counts. The influx of Omicron in December 2021 had an exponential increase in the monthly average of observed changes in immune epitopes (**Fig. 4B**). When examining antiviral resistance markers toward reduction of IC50 for Ensovibep (ACE-2-binding inhibitor), Nirmatrelvir (protease inhibitor) and Remdesivir (viral polymerase inhibitor) within the different VOC, we identified spurious occurrences of resistance mutations against Ensovibep within the spike gene and resistance mutations against Nirmatrelvir and Remdesivir within ORF1ab were identified in isolates of Iota, Delta and Omicron, although not Alpha (**Fig. 4A & 4B**). There was a consistent count of T-cell epitope mutations per virus throughout the second year. However, antibody resistance mutations exhibited a non-significant increase coinciding with the introduction of the Delta variant in June 2021, and a significant increase in both types of immune resistance mutations after December 2021, driven by Omicron’s arrival (**Fig. 4C**). Iota had a wider range of epitope changes per virus depending on the time it was sampled, necessitating the use of a sampling time controlled linear model to assess the difference in the number of epitope mutations for the different VOC. Alpha, Iota and Delta did not significantly differ in T-cell epitope changes (p-value: 0.3-0.5), while all variants exhibited statistically less changes than Omicron (p-values < 1.2 x10^-137^). All VOC had statistically different numbers of antibody epitope changes with each other (p-values < 3×10^-7^), Alpha having the lowest number and Omicron having the most. To simultaneously consider the number of epitopes and the overlap in epitope changes, the profile of T-cell epitope mutations and antibody escape mutations were both decomposed into two axes of variation via PCA. The strongest separation between variants was between Omicron and the rest of the variants and explains roughly 45-54% of the variance in immune epitope changes. Both Iota and Delta were separated from the ancestral viruses and Alpha in the second principal component axis (13-15% of variance). Delta’s and Iota epitope variability were minor relative to Omicron, but still separable from ancestral viruses, unlike Alpha (**Fig. 4D**).

**Figure 4:**
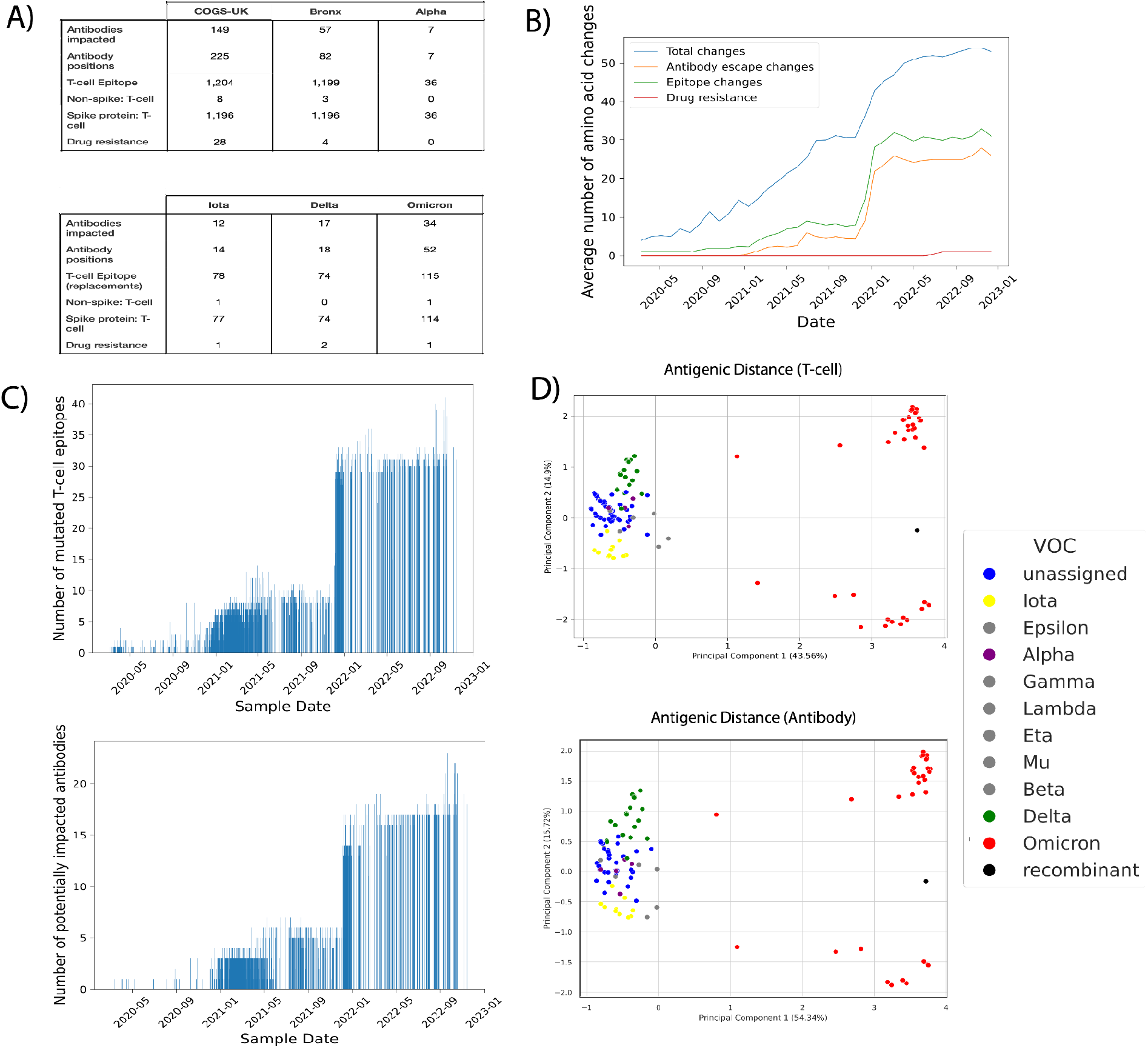
Population-level count of epitope mutations suggests Bronx Alpha VOC has the lowest epitope diversity and Bronx Delta VOC has relatively few novel T-cell epitopes compared to Iota. **A)** Population counts of unique antibody resistance mutations, the antibodies they impact, T-cell epitope mutations, and drug resistance markers in the entire COGS-UK database, the Bronx sequences, or the major VOC observed in the Bronx. **B)** Plot of monthly sampling rates of total amino acids and changes in antigenic sites. **C)** Plots of the number of known T-cell epitopes and antibodies impacted by amino acid changes per sampled virus over time. **D)** Two PCA plots depicting the relationship of Bronx viral isolates by their T-cell or antibody epitope change profiles, with colors representing different VOC/VOI.

## Discussion

A major goal of SARS-CoV-2 surveillance is to uncover generalizable trends in the rates of COVID-19 cases and hospitalizations to inform public health strategies, yet it remains unclear what level of geographical granularity is most informative. Here, we show that in the Bronx, deviations from national trends were evident in the second year of the pandemic. These deviations reveal VOC interactions and patient outcomes that differ from those observed at larger geographic scales and highlight the need for local surveillance. We defined the transmission dynamics of Delta, Iota, and Alpha, which were the major sampled variants during the spring and summer of the second year of the pandemic. We highlight three specific features of the SARS-CoV-2 pandemic in the second year that differentiate the Bronx from the U.S.A: the dominance of Iota in place of the Alpha variant in early 2021, lower-than-average hospitalizations associated with the Delta variant in the same year, and higher-than-average hospitalizations associated with the introduction of the Omicron variant.

One feature of distinct COVID-19 VOC dynamics in the Bronx was the dominance of the VOC Iota at a time when Alpha was the globally dominant strain at the time of co-circulation and was described to have superior binding efficiency to human ACE-2 receptors (Carabelli et al. 2023). We explored whether Iota prevented the dominance of Alpha in the Bronx. We determined that Iota and Alpha had similar mutational fitness profiles and developed a SIR model to assess differences in infection in Iota and Alpha. While Alpha exhibited a higher probability of infection than Iota, Iota had pre-existing residency in the Bronx and a lower rate of recovery, resulting in a similar measure of transmission fitness. Additionally, Iota possessed a broader array of immune escape mechanisms due to strain variability and others have noted greater resistance (higher IC50) to vaccinated sera than Alpha (Annavajhala et al. 2021 & Choi et al. 2021). Our findings also indicate there is greater variability in T-cell epitopes in Iota Bronx strains than previously reported in Iota. Limited epidemiological studies suggest these two VOCs had equivalent immune escape capacities with equal vaccine breakthrough infection rates (Duerr et al. 2021). Other studies have suggested that Alpha has had very few antigenic escape mechanisms which supports our observations in the Bronx (García-Beltrán et al. 2021, Brown et al. 2021 & Newman et al. 2022). Overall, the epitope observations in conjunction with the SIR model results suggest that there was a trade-off between decreased γ in Iota infections and an increased β in Alpha infections resulting in an equivalent fitness (**Table 1**), as predicted by the PyR_0_ mutation fitness model (**Fig. 2B**).

Our local study encompasses all reported, high-quality, sequences from the Bronx, providing a more comprehensive perspective on the dynamics of Alpha and Iota in this region. Our observations support an equal overall fitness between Alpha and Iota interactions with the Bronx population, with the pre-existing establishment of Iota in the Bronx preventing the spread of Alpha. The aggregated data from the U.S.A. suggests, on average, that the dynamics have played out differently elsewhere because of Alpha’s known mechanisms of increased transmission speed via more efficient ACE2 binding (Carabelli et al. 2023).

We next turned to the distinctive pattern in the Bronx’s case rates, where the Delta peak in the summer of 2021 falls below the 99% confidence interval for mean Bronx cases despite a higher than average community viral load, as revealed by wastewater PCR testing for the SARS-CoV-2 N gene (**Fig. 1A**). Given that there was little variant sub-diversity at this point and continued detection of the virus, hospitalizations in the Bronx are below average during the summer of 2021, diverging sharply from the national aggregate, which remains above average during the same period (**Fig. 1B**). Together, these observations support a hypothesis suggesting that fewer people were symptomatic or seeking treatment because their cases were less severe, resulting in lower-than-average COVID-19 cases and hospitalizations despite above-average SARS-CoV-2 wastewater detection. The observation that Delta does not have higher hospitalizations per case goes against the conventional argument that Delta was a more virulent VOC. Hospitalization and case data instead suggest that the community-level impact of hospitalization per case of Delta was equivalent to the preceding peak of mainly Iota and Alpha cases.

In contrast with Alpha and Iota, Delta was infecting hosts with greater immunity against SARS-CoV-2, as the population became more vaccinated or developed natural immunity from prior infections. Immunity in the hosts would reduce severity and possibly reduce its spread amongst the Bronx patient population. Despite these disadvantageous conditions, Delta had advantages over other VOCs such as an increased mutational fitness, and a slight increase in antibody resistance per virus. Delta displayed fewer novel T-cell epitope mutations compared to the Iota variant, which was surprising given it was an ancestral strain. T-cell activation is important in mounting a fast immune response and prevention of severe disease (Julia Maret Hermens and Can Keşmir 2023); the lower community-level virulence of Delta is thus possibly linked to a lack of new immune escape mechanisms relative to the preceding strains the Bronx patients were exposed to. We propose that Delta’s advantages in terms of mutations (**fig. 2A & 2B**) and antibody resistance (**fig. 3C & 3D**) might be counteracted by higher pre-existing immunity within the community due to the dominance of the Iota VOI.

Modeling of transmission dynamics supports the notion that higher pre-existing immunity of the Bronx population reduced the community virulence of the more transmissible Delta VOC. Here, our two SIR models included the assumption that there was a higher amount of pre-existing immunity by incorporating a recovered fraction that represents the reported cumulative number of infections at the start of the simulation, which is biologically reasonable given that immunity shouldn’t wane within a year (Stein et al. 2023) given that there were few novel epitope mutations. Delta’s higher probability of recovery from infection (γ) complements the genomic observation of equivalent numbers of T-cell epitope mutations in Delta and Iota (**Table 1 & Fig. 4A & 4D**). The probability of recovery is inversely proportional to the length of time to recover and acquire immunity, and the larger γ of Delta suggests that a quick recovery time from Delta infections dampened its overall impact. Together, our work, in concert with that of others, indicates the importance of prior immunity impacting the recovery rate.

Finally, we focused on differences in hospitalizations in the Bronx during Omicron’s prevalence, which were higher than the local average (**Fig. 1C**). State-wide hospitalizations due to COVID-19 peaks were normalized and overlapped in December 2021 and were above each time series average too, suggesting a period of uniformity nationwide. Within the Bronx there was a significant correlation between Bronx Omicron cases and hospitalizations associated with the first introduction of SARS-CoV-2, suggesting similarities to a naive introduction. Evidence from Hong Kong suggests that Omicron sublineage BA.2 displays a hospitalization frequency similar to that of first-wave variants (Mefsin et al. 2022) which aligns with our autocorrelation analysis based on the Bronx population (**Supp. Fig. 1**).

In conclusion, our integrated analysis of SARS-CoV-2 and COVID-19 cases and hospitalization trends in the Bronx over the second year of the pandemic underscores the critical importance of localized studies in understanding the intricacies of viral dynamics. The deviations observed in the Bronx from national trends, such as the dominance of the Iota variant over Alpha in early 2021 and the unexpected reduction in hospitalizations associated with the Delta variant, reveal the need for a more granular examination of the pandemic’s impact in local areas. The distinctive pattern in case rates, the discrepancy between diagnosed cases and community viral load, and the complex interplay between hospitalization trends and viral variants add layers of complexity to the local dynamics. The Delta variant’s unique challenges to the Bronx, including a higher mutational fitness and antibody resistance per virus, were mitigated by factors such as pre-existing immunity within the community. The observed dynamics between Alpha and Iota in the Bronx may not universally apply beyond the region, emphasizing the importance of localized studies in unraveling the complexities of the pandemic. Overall, our analysis highlights localized factors influencing viral dynamics that may be missed in larger, countrywide, or global studies, providing a foundation for tailored local public health interventions.

## Methods

### Public Health Data

All Bronx COVID-19 epidemiological data, which includes: diagnosed cases, hospitalizations, deaths, and testing rates, comes from the NYC Department of Health and Mental Hygiene (https://github.com/nychealth). All community health measures for COVID-19 are defined by the most up-to-date definitions by the CDC’s Council of State and Territorial Epidemiologists (“Coronavirus Disease 2019 (COVID-19) 2021 Case Definition | CDC” 2021). Wastewater community SARS-CoV-2 viral loads are based on qPCR copies of the viral nucleocapsid gene per liter of water sampled from two locations in the Bronx: Hunts Point and Ward’s Island (https://data.cityofnewyork.us/Health/). Data for trends in the estimated VOC cases in the United States was curated from GISAID: https://gisaid.org, John Hopkins COVID-19 Data Repository: https://github.com/CSSEGISandData/COVID-19, and obtained from Covariants https://covariants.org/cases, while the U.S.A aggregate hospitalizations and state hospitalizations were obtained from the CDC’s COVID-NET database: https://www.cdc.gov/coronavirus/2019-ncov/covid-data/covid-net/purpose-methods.html.

### Time Series & Epidemiological Statistics

All scripts were custom and written in Python. Stationarity, a behavior describing the independence of the variance and mean from time, was assessed to measure a constant baseline. The Augmented Dicker-Fulley test checked for stationary behavior and normality assumption was assessed with the Shapiro-Wilk test and visually checked with Q-Q plots (**Supp. Fig. 1**). All time series were considered stationary, meaning they have a constant mean and variance over time and normalizing the time-series means were used to compare the relative magnitude of cases or hospitalizations.

The autocorrelation analysis was conducted using the pandas autocorrelation_plot function in Python. This function generates a plot of the autocorrelation coefficients of the time series, which can be used to identify any patterns or trends in the data over time. The autocorrelation coefficient is a measure of the correlation between a variable and its lagged values.

The autocorrelation coefficient of lag k can be calculated using the following formula:

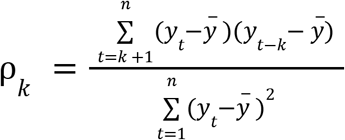

Where y is the time series variable, n is the sample size, and k is the lag. To calculate the confidence interval, the function uses the formula:

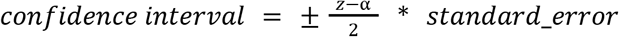

Where 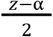 is the critical value for the normal distribution at the 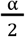 significance level (where

alpha is the significance level), and standard_error is the standard error of the autocorrelation estimate. The standard error is estimated as:

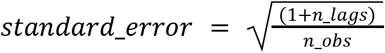

where n_lags is the number of lags being plotted and n_obs is the total number of observations in the time series.

To measure the correlation between daily hospitalizations and daily cases, values ranging from January 1st, 2021 to October 31st, 2021 were selected for the periods where Iota dominated and the peak in cases where only Delta was detected (no Omicron detected). The MASS package was used to create a robust linear model with an interaction term for the dominating VOC and the cases and the F-test was used to test the significance of that term’s influence on the daily hospitalizations: *H* ∼ β_0_ + β_1_*C* + β_2_*C* * *V*, where *H* = hospitalizations, *C*= cases, *V* = dominating VOC.

### Genomic Data

The Global Initiative on Sharing All Influenza Data (GISAID) was used to collect 2,792 Bronx SARS-CoV-2 sequences and associated metadata. The GISAID database (Shu and McCauley 2017) is an open-access platform that facilitates the sharing of viral sequences and associated metadata for research and public health purposes. The sequences were filtered for high-quality sequences based on GISAID’s quality control criteria.

### Epidemiological Modeling

Kernel density smoothed relative frequency of sampled cases by each VOC over time was used to estimate the proportion of cases contributed by each VOC over time. The resultant data was then used to fit independent epidemiological models to infer transmission and recovery parameters.

A linear regression model was used to find a constant short-term estimate for the force of infection: *λ* = *c* * β, where *λ* = force of infection, c = contact rate, and β = probability of transmission. We estimated *λ* for the different variants by regressing cumulative cases: C(t), with new cases: I(t), given the linear relation: *C*(*t*) = *λ* * *I*(*t*) (Md. Kamrujjaman et al. 2022).

Both a dynamic and stochastic SIR model were implemented to estimate the transmission rate: *c*β, recovery rate: *γ* and the reproduction number: *R*_0_ of each VOC observed during the second year. It assumes a virus has a susceptible host: *S*(*t*) and becomes infected: *I* (*t*), with some rate that is the product of the probability of infection: β and the contact rate: *c*. The host recovers and acquires immunity: *R*(*t*), at rate: *γ*, and is assumed to have lasting immunity for the duration of the simulation which is under a year (Stein et al. 2023). SARS-CoV-2 spread is dependent on population density so the transmission parameter is divided by the size of the total population: *N* = *S*_0_ + *I*_0_ + *R*_0_ . The following differential equations govern these dynamics: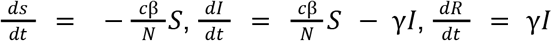. The dynamic model is solved using the DeSolve package in R and utilizes the least squares method to find the best fitting parameters to the smoothed case data (Md. Kamrujjaman et al. 2022).

In the Stochastic model, we use stochastic differential equations which model the fluxes between states as counting processes:

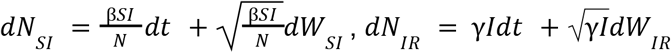, where dW is an independent normal distribution: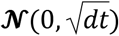, which can be thought of as a noise term in the differential equation. This gives us the following stochastic differential equations: 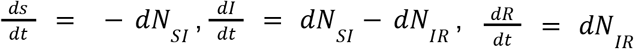. We use the Euler-Maruyama method to solve these equations via a custom R script (Jürgen vom Scheidt 1994) and fit the mean trajectory via the least square method (Md. Kamrujjaman et al. 2022).

### Nextstrain analyses

The default Nextstrain pipeline (Hadfield et al.) was used for molecular epidemiological analysis of the 2,792 Bronx sequences. The pipeline involved several steps, including alignment, phylogenetic inference, and visualization. The fasta sequences were aligned using MAFFT (Katoh 2002), Maximum likelihood phylogenetic inference was then performed using IQ-TREE (Nguyen et al. 2014), and the resulting tree was visualized using the auspice web application (Hadfield et al.). The pipeline was run using the Snakemake workflow management system, which is facilitated for reproducibility and scalability. The time-scaled phylogenetic trees were regressed with the number of mutations and the mutational fitness. PyRo, a hierarchical Bayesian multinomial logistic regression model, was used as a part of the Nextstrain workflow to estimate the relative fitness of each virus by the combination of mutations associated with its continued spread (Obermeyer et al. 2022).

### Quantifying epitope mutations and drug resistance mutations

We utilized the alignments generated by the Nextstrain pipeline to annotate single amino acid changes relative to the Wuhan reference. To further investigate the potential implications of these amino acid changes, we utilized the COG-UK Mutation Explorer: https://sars2.cvr.gla.ac.uk/cog-uk/, a comprehensive SARS-CoV-2 database that focuses on observed amino acid replacements with antigenic roles in the human immune response. This database contains over 2 million genome sequences specifically for UK SARS-CoV-2 data, which is curated and analyzed to track mutations of immunological importance that are accumulating in current variants of concern and variants of interest. These mutations have the potential to alter the neutralizing activity of monoclonal antibodies, convalescent sera, and vaccines, as well as changes in epitopes recognized by T cells, including those with reduced T cell binding. The database also includes mutations that have been shown to confer SARS-CoV-2 resistance to antiviral drugs (Wright et al. 2022). All analyses were made using custom Python scripts to count antibody resistance mutations, the antibodies impacted by them, the number of mutations in t-cell epitopes, and drug resistance-associated mutations. Single amino acid polymorphisms (**SAAPs**) in the local Bronx sequences were called using the Nextclade pipeline and cross-referenced with the SAAPs in the COG-UK Mutation Explorer database to create a set of immune epitope changes for each sampled Bronx virus. A PCA was used to cluster antigenic relationships between individual viruses based on the immune epitope changes they share. To test for quantitative differences in epitopes, an ordinarily least squares model was used comparing each viruses contribution to the variation in epitope mutations (*E*) over time. The following formula describes the model:

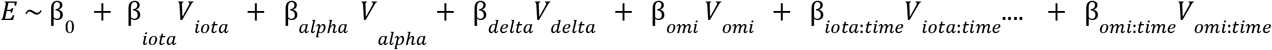

The β values for each virus were compared pairwise via an F-test to obtain statistical significance.

## Data Availability

The source code used for analysis and figure generation, is hosted on Github at https://github.com/kellylab/SARS-CoV-2-surveillence.git.

https://github.com/nychealth

https://data.cityofnewyork.us/Health/

https://gisaid.org

https://github.com/CSSEGISandData/COVID-19

https://covariants.org/cases

https://www.cdc.gov/coronavirus/2019-ncov/covid-data/covid-net/purpose-methods.html

https://sars2.cvr.gla.ac.uk/cog-uk/

## Acknowledgements

We extend our gratitude to NextStrain, GISAID, and all the laboratories that have contributed SARS-CoV-2 sequences for public access. Our heartfelt thanks go to the healthcare workers and patients of the Montefiore Healthcare System and other health systems in the Bronx who have participated in The New York City Department of Health and Mental Hygiene’s COVID-19 public health data collection efforts. Additionally, we express appreciation to the Centers for Disease Control, the New York State Department of Health, and the Department of Health and Mental Hygiene for their centralization and facilitation of public access to COVID-19 public health data for the Bronx, NY, and the USA.

## Author contributions

Study design: L.K., J.P.D, R.F. Data analysis: L.K., R.F., A.G. Wrote paper: R.F.. Edited paper: all authors.

## Supplementary materials

Figs. S1 to S4

