## Supplemental figures for "Community level variability in Bronx COVID-19 hospitalizations associated with differing viral variant adaptive strategies during the second year of the pandemic"

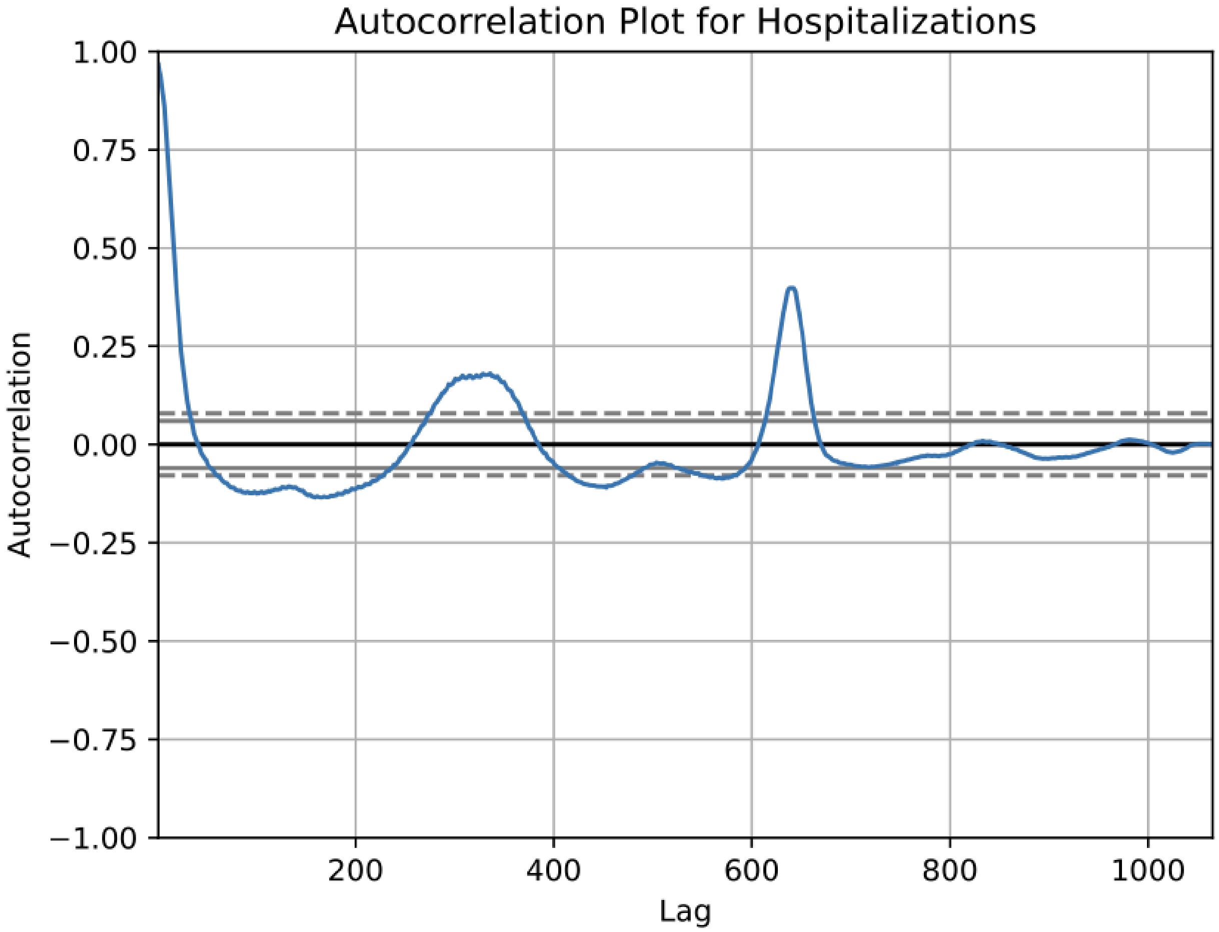


**Supplemental figure 1:** Autocorrelation of hospitalizations due to COVID-19 indicate 3 statistically correlated peaks across different days (lag), as they are correlation values above the 99% convince interval (dotted line). The First peak in winter 2020 is at the beginning, then winter 2021 and winter 2022 as the winter peaks continue they appear to increase in magnitude. Omicron was first introduced in 2021 lagging more than 600 days after the initial peak in hospitalizations.

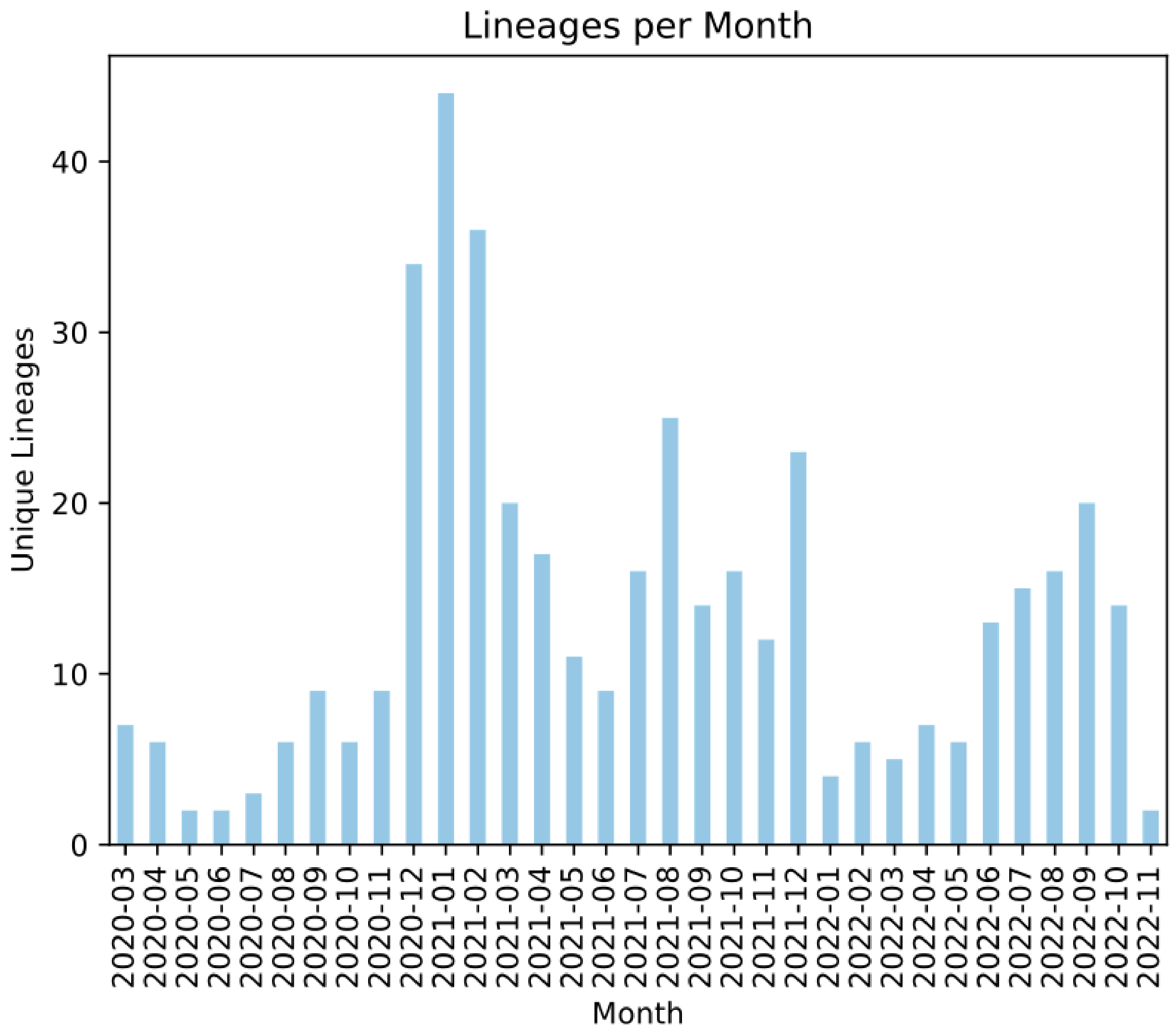


**Supplemental figure 2:** Unique number of pangolin lineages per month. Pangolin lineages are geographic dependent clustering of closely related viruses and represent a granular view of viral diversity. The peak in lineage diversity was observed in January 2021 during the initial introduction of vaccines in the Bronx.

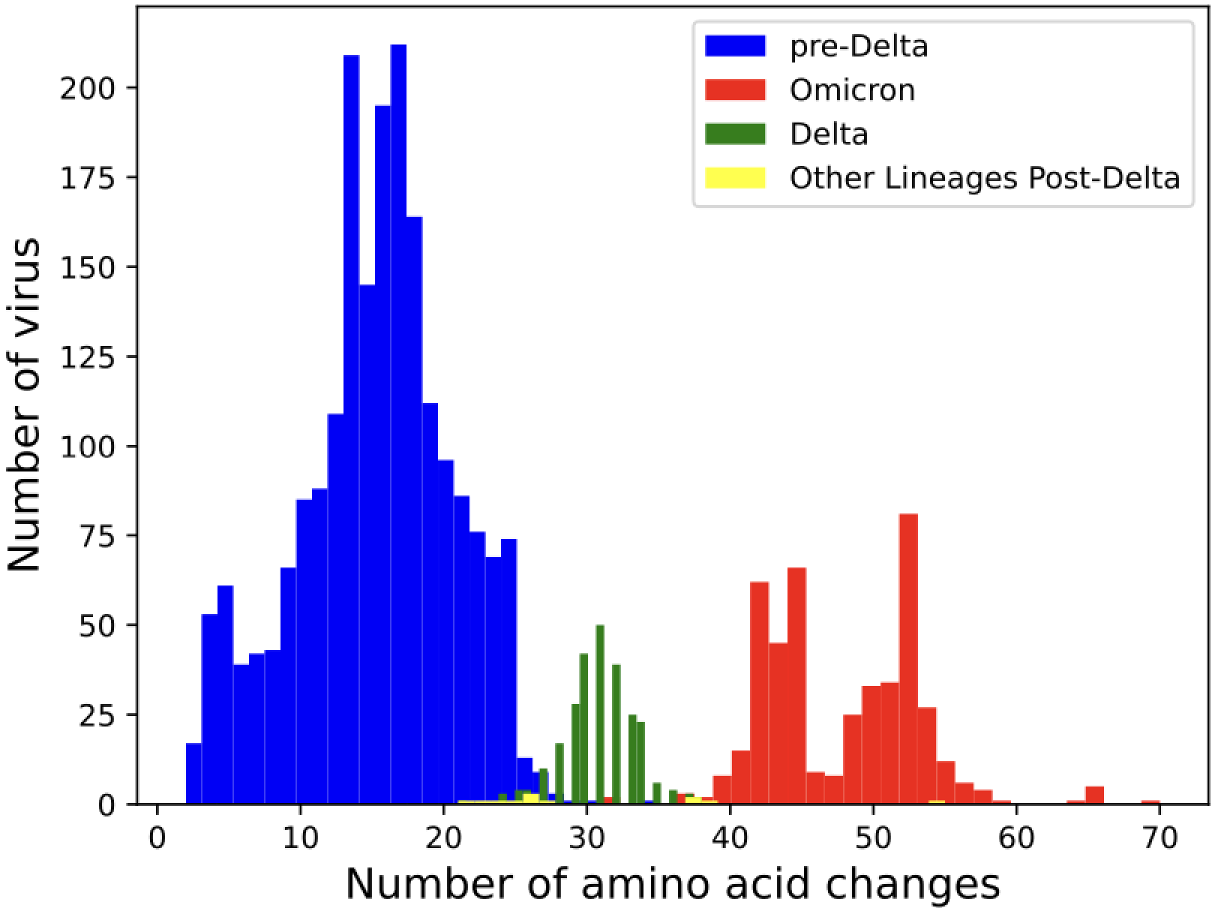


**Supplemental figure 3:** Histogram of non-synonmous mutations (single amino acid changes) of Omicron, Delta and Pre-Delta sequences form roughly three statistically different distributions by ANOVA (p-value < 0.001).

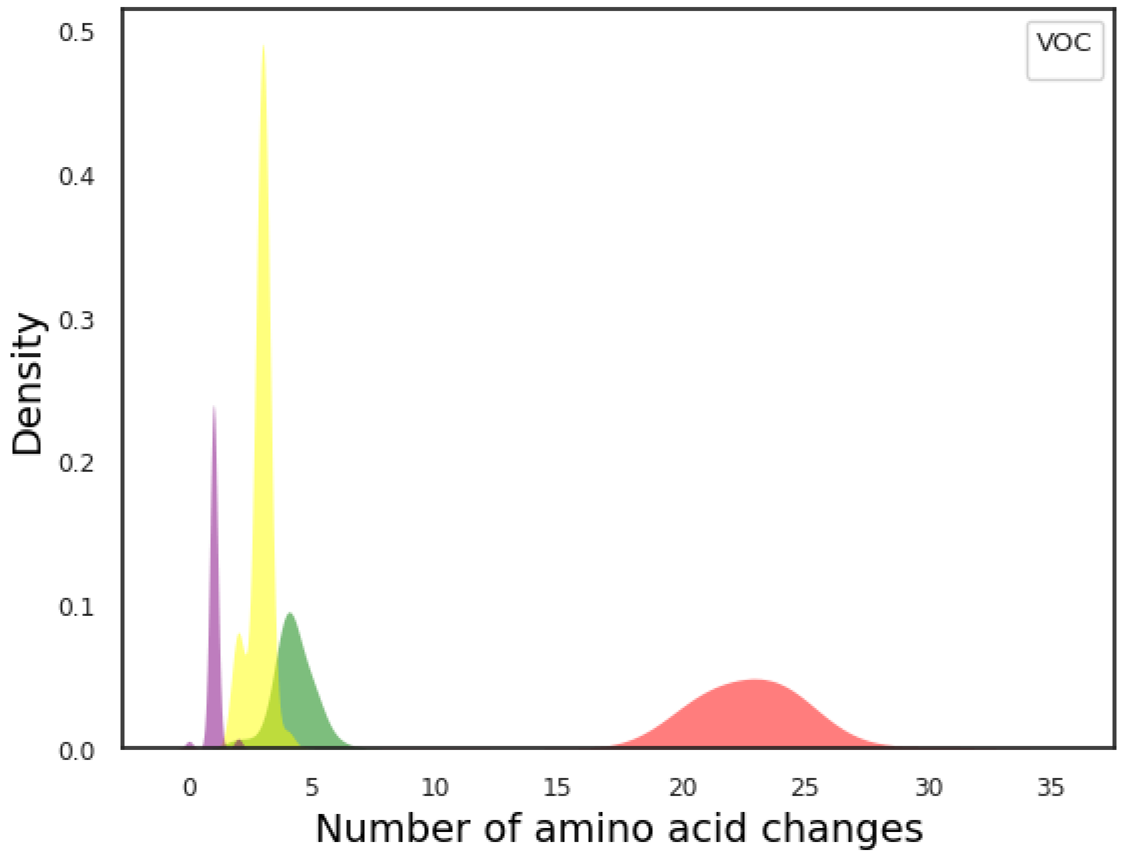


Supplemental figure 4: Density plot of antibody impacting mutations of Bronx Omicron, Delta, Iota and Alpha sequences form four statistically different distributions by a time-controlled linear model (p-value < 0.001).

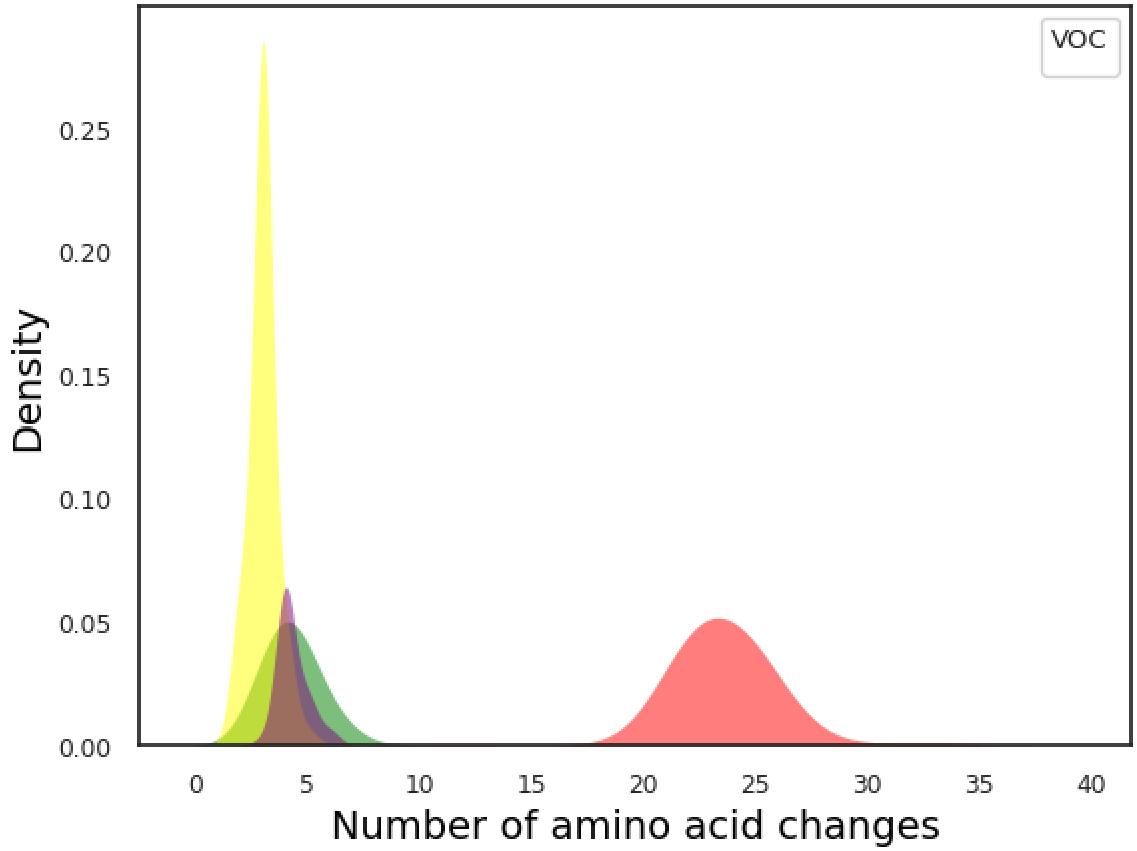


Supplemental figure 5: Density plot of t-cell epitope mutations of Bronx Omicron, Delta, Iota and Alpha sequences form two statistically different distributions by a time-controlled linear model (p-value < 0.001).
